# Brain correlates of suicide attempt in 18,925 participants across 18 international cohorts

**DOI:** 10.1101/2020.05.06.20090191

**Authors:** Adrian I. Campos, Paul M. Thompson, Dick J. Veltman, Elena Pozzi, Laura S. van Veltzen, Neda Jahanshad, Mark J. Adams, Bernhard T. Baune, Klaus Berger, Katharina Brosch, Robin Bülow, Colm G. Connolly, Udo Dannlowski, Christopher G. Davey, Greig I. de Zubicaray, Danai Dima, Tracy Erwin-Grabner, Jennifer W. Evans, Cynthia H.Y. Fu, Ian H. Gotlib, Roberto Goya-Maldonado, Hans J. Grabe, Dominik Grotegerd, Matthew A. Harris, Ben J. Harrison, Sean N. Hatton, Marco Hermesdorf, Ian B. Hickie, Tiffany C. Ho, Tilo Kircher, Axel Krug, Jim Lagopoulos, Hannah Lemke, Katie McMahon, Frank P. MacMaster, Nicholas G. Martin, Andrew M. McIntosh, Sarah E. Medland, Susanne Meinert, Tina Meller, Igor Nenadic, Nils Opel, Ronny Redlich, Liesbeth Reneman, Jonathan Repple, Matthew D. Sacchet, Simon Schmitt, Anouk Schrantee, Kang Sim, Aditya Singh, Frederike Stein, Lachlan T. Strike, Nic J.A. van der Wee, Steven J.A. van der Werff, Henry Völzke, Lena Waltemate, Heather C. Whalley, Katharina Wittfeld, Margaret J. Wright, Tony T. Yang, Carlos A. Zarate, Lianne Schmaal, Miguel E. Rentería, for the ENIGMA-MDD working group

**Author notes:** These authors jointly supervised this study (Lianne Schmaal, Miguel E. Rentería). Correspondence: Miguel E. Rentería.

## Abstract

**Background:** Suicidal behavior is highly heterogeneous and complex. A better understanding of its biological substrates and mechanisms could inform the design of more effective suicide prevention and intervention strategies. Neuroimaging studies of suicidality have so far been conducted in small samples, prone to biases and false-positive associations, yielding inconsistent results. The ENIGMA-MDD working group aims to address the issues of poor replicability and comparability by coordinating harmonized analyses across neuroimaging studies of major depressive disorder and related phenotypes, including suicidal behavior.

**Methods:** Here, we pool data from eighteen international cohorts with neuroimaging and clinical measurements in 18,925 participants (12,477 healthy controls and 6,448 people with depression, of whom 694 had attempted suicide). We compare regional cortical thickness and surface area, and measures of subcortical, lateral ventricular and intracranial volumes between suicide attempters, clinical controls (non-attempters with depression) and healthy controls.

**Findings:** We identified 25 regions of interest with statistically significant (FDR<0.05) differences between groups. *Post-hoc* examinations identified neuroimaging markers associated with suicide attempt over and above the effects of depression, including smaller volumes of the left and right thalamus and the right pallidum, and lower surface area of the left inferior parietal lobe.

**Interpretation:** This study addresses the lack of replicability and consistency in several previously published neuroimaging studies of suicide attempt, and further demonstrates the need for well-powered samples and collaborative efforts to avoid reaching biased or misleading conclusions. Our results highlight the potential involvement of the thalamus, a structure viewed historically as a passive gateway in the brain, and the pallidum, a region linked to reward response and positive affect. Future functional and connectivity studies of suicidality may focus on understanding how these regions relate to the neurobiological mechanisms of suicide attempt risk.

## INTRODUCTION

Suicide is a leading cause of death worldwide and is a considerable health concern in both developed and developing countries^1^. While region and country-specific estimates vary, the global average prevalence for suicide is estimated to be about 10.6 deaths per 100,000^2^. Suicide attempts outnumber actual suicides by twenty to thirty-fold,^3,4^ which further increases the economic and social burden of suicidal behavior^5^.

Suicidal behavior is more common in people living with mental illness^6–8^. In fact, for a long time, suicidality was conceptualized as a symptom inherent to certain conditions, in particular, major depressive disorder (MDD). However, a recent report from the U.S. showed that while suicide rates are increasing, approximately 54% of suicide decedents in 2015 had no diagnosis of any mental disorder^9,10^. On the whole, a better understanding of suicidality, in terms of its underlying mechanisms, could help identify individuals at increased risk of engaging in suicidal behaviors and inform better interventions.

Non-invasive neuroimaging technologies, such as magnetic resonance imaging (MRI), allow brain structure and function to be studied *in vivo*.^11^ The analysis of brain morphometry and neuroanatomical differences, on average, between individuals with mental illness and healthy controls has already proven useful in a range of conditions such as MDD^12^, bipolar disorder^13^ and schizophrenia^14^. Similar approaches have been used to study suicidality, albeit in small samples. Briefly, several studies have reported lower grey matter volume and cortical thickness in the frontal, prefrontal,^15–23^ and temporal ^16–20,24–26^ lobes associated with suicidality.

Nonetheless, small samples and heterogeneous analysis methods have led to a lack of replicability and inconsistent results.^27,28^ The ENIGMA-MDD working group aims to address issues of poor replicability and comparability in neuroimaging studies by coordinating harmonized analyses of MDD and related phenotypes, including suicidal behavior. In the most recent meta-analysis of subcortical brain volumes conducted by our working group with a sample size of 3,097, we did not detect any significant morphological differences associated with suicidality independently of depression diagnosis^11^.

Here we present the largest and most comprehensive neuroimaging study of suicide attempt to date. We perform a pooled mega-analysis of subcortical volumes and regional cortical surface area and thickness, using linear mixed model regressions in a sample of 18,925 subjects from eighteen cohorts from around the world. We aim to shed light on the neural circuits that underlie suicidal behavior by comparing brain morphometry between MDD cases with a history of suicide attempt versus those without, as well as versus healthy controls.

## METHODS

### Samples

We analyzed pooled data (mega-analysis) across seventeen ENIGMA-MDD working group cohorts with clinical and neuroimaging data available for participants fulfilling MDD criteria^29^ (N=2,533) and healthy controls (N=4,066), and participants from the UK Biobank (N=12,326). We defined three groups: *suicide attempters* (SA), *clinical controls* (CC), that is, participants with depression and no history of suicide attempt, and *healthy controls* (HC). Descriptive statistics for each sample are listed in **Table 1** and **Supplementary Table S1**. Each cohort assessed depression status and history of a suicide attempt based on available clinical information. In the UK Biobank, lifetime depression status (N=3,633) and lifetime suicide attempt (N=322) were ascertained using the Composite International Diagnostic Interview (CIDI). Participants with no history of depression or suicide attempt (N=8,411) were defined as healthy controls. Detailed information on the diagnostic instruments used to determine MDD and suicidality, as well as exclusion criteria, are available in **Supplementary Table S2**. The combined sample comprised 12,477 healthy controls and 6,448 participants with a lifetime depression diagnosis. Within the depression group, 694 participants reported at least one suicide attempt. All sites obtained approval from their local institutional ethics committees and review boards to participate in this study, and all participants provided informed consent at their local recruitment institution.

**Table 1.** Demographics and clinical measures across studied groups

|  | HC | CC | SA |
| --- | --- | --- | --- |
| Total N (%) | 15,269 (71) | 5,557 (26) | 694 (3) |
| Females N (%) | 7,637 (50) | 3,642 (66) | 466 (67) |
| Males N (%) | 7,632 (50) | 1,915 (34) | 228 (33) |
| Age mean (sd) | 57.6 (14.8) | 53.2 (15.4) | 49.2 (16.3) |
| BDI mean (sd)<sup>‡</sup><sup>Ω</sup> | 3.6 (4.0) | 18 (10.9) | 23 (11.8) |
| HDRS mean (sd)<sup>‡</sup><sup>Ω</sup> | 1.3 (2.0) | 11.0 (6.9) | 13.8 (6.9) |
| Depression age of onset<sup>‡</sup> | NA | 29.8 (14.3) | 23.3 (14.1) |
| Antidepressant use %<sup>‡</sup> | 0.1% | 13% | 11% |
| Depression recurrence<sup>‡</sup> % | NA | 21% | 36% |

HC- Healthy controls; CC- Clinical controls; SA - Suicide attempters.
BDI- Beck Depression Inventory; HDRS- Hamilton Depression Rating Scale
<sup>‡</sup>Data available only for a subset of the sample. <sup>Ω</sup>Sum-score excluding the suicidality item.

### Image processing and analysis

T1-weighted MRI structural brain scans were acquired and analyzed locally at each site using the validated and automated segmentation software *FreeSurfer*^30^ (available at http://surfer.nmr.mgh.harvard.edu/). Image acquisition parameters and software versions and descriptions are detailed in **Supplementary Table S2**. The segmentation of cortical and subcortical phenotypes was visually inspected for accuracy following standardized protocols designed to facilitate harmonized image analysis across multiple sites (http://enigma.ini.usc.edu/protocols/imaging-protocols/). Within each cohort, histogram plots were generated for each region to examine the distribution of each volumetric variable. Measures were visually verified for accuracy and excluded if they were not properly segmented. Within-cohort outliers (defined as measurement greater than three standard deviations away from the mean) were excluded from the analysis. We examined five *global* brain measures (intracranial volume, total surface area of the left and right hemispheres and mean cortical thickness of the left and right hemispheres), 16 subcortical brain volume measures, and cortical surface area and thickness measures for 68 brain regions of interest as defined by the Desikan-Killiany Atlas.

### Ascertainment of suicide attempt history

Suicide is the act of intentionally ending one’s life. In this study, a suicide attempt was defined as any self-harm act with the intent to die. Attempt severity was not assessed due to a lack of information in individual studies. A description of how suicidality was measured in each independent site is available in **Supplementary Table S2**. Cohorts also provided (where available) information on i) whether participants have used antidepressants, ii) depression severity, coded either as the Hamilton Depression Rating Scale (HDRS) score excluding the suicide item, or the number of DSM-IV MDD criteria endorseed (ranging from 0 to 9), iii) age of depression onset and iv) whether depression was recurrent or a single episode.

### Statistical analyses

#### Linear mixed-effects models

Statistical analyses were performed in R v3.6.1 using the statistical package *lme4*. To cope with the relatively low prevalence of suicide attempts within each cohort, we conducted a mega-analysis. Linear mixed-effects models were used to account for site variation (with a random intercept for scan site) while correcting for desired covariates as fixed effects. We modeled each regional measure as an outcome while using an indicator variable per group of interest: healthy controls (HC), MDD patients with no suicide attempt history (clinical controls; CC), and MDD patients with attempt history (SA). All models adjusted data for age and sex, while surface area and volumetric analyses also adjusted for ICV (except when ICV was the measure of interest). Main effects of groups (i.e., differences between groups of healthy controls, clinical controls, and clinical attempters) were identified by performing a type II analysis of variance (*F*-test) over the fitted linear mixed-effects model described above. We conducted follow-up (*post-hoc*) analyses to assess whether the effects were driven by suicide attempt over and above MDD status. To this end, regions of interest were compared between suicide attempters and clinical controls, and between suicide attempters and healthy controls. Finally, we conducted sensitivity analyses to assess the effects of severity, recurrence, and age of onset of depression, and history of antidepressant use on the observed associations. To achieve this, we repeated the *post-hoc* analyses of the four regions showing evidence of association with suicidality, including additional covariates one at a time.

#### Statistical significance definition

We corrected for multiple comparisons using a false discovery rate (FDR) procedure^31^ for each set of morphometry measures separately: subcortical volumes, cortical thickness, and cortical surface area. The significance threshold to define regions of interest (ROI) for *post-hoc* analyses was set at FDR *p*-value <0 05. Post-hoc regressions comparing attempters to clinical and healthy controls used a matrix spectral decomposition to identify the number of effective variables ^32,33^ coupled with Bonferroni to keep the type I error rate at 5%. In this manuscript, a *significant* result survived multiple testing corrections (p<Bonferroni corrected threshold), whereas a *nominally significant* result was only significant before correction (p<0.05).

## RESULTS

### Suicide attempt prevalence and sample demographics

Details on the sample size and age and sex composition of each cohort included in this analysis are summarized in **Table 1 and Supplementary Table S1**. Notably, not all cohorts had cases of suicide attempt, but still contributed data to the healthy or clinical control groups. The pooled mean age was 56.22 years old, with a standard deviation of 15.17 years. Differences in age and sex composition across cohorts were detected (**Supplementary Table S1**) and used as covariates for all the analyses. The total sample size comprised 18,925 subjects, of which 3.67% (N=694) had at least one past suicide attempt. Furthermore, 30.04% (N=5,574) of the total sample was diagnosed with depression but did not report a previous suicide attempt. Methodological differences (e.g., scanner used or different parameters for the scan) between participating cohorts are listed in **Supplementary Table S2**. We used mixed-effects models (**see Methods**) to account for systematic site differences that could bias observed associations.

### Subcortical volumetric measures

The thalamus (right and left), right pallidum, and total ICV exhibited a statistically significant group effect (i.e., any difference between healthy controls, clinical controls, or suicide attempters) after correcting for multiple comparisons. The left pallidum and the right nucleus accumbens showed a nominally significant difference but did not survive correction for multiple comparisons (**Supplementary Table S3**). Depressed attempters exhibited smaller volumes in the left and right thalamus and right pallidum compared to both clinical (Cohen’s d=−0.13, −0.14 and −0.12, respectively) and healthy controls (**Figure 1**). Depressed attempters also exhibited smaller ICV compared to healthy controls (Cohen’s d =−0.13). This association did not reach significance, when comparing attempters and clinical controls, after accounting for multiple testing. None of the regions with a significant group effect showed a significant difference when comparing clinical to healthy controls (**Table 2**).

**Figure 1.**
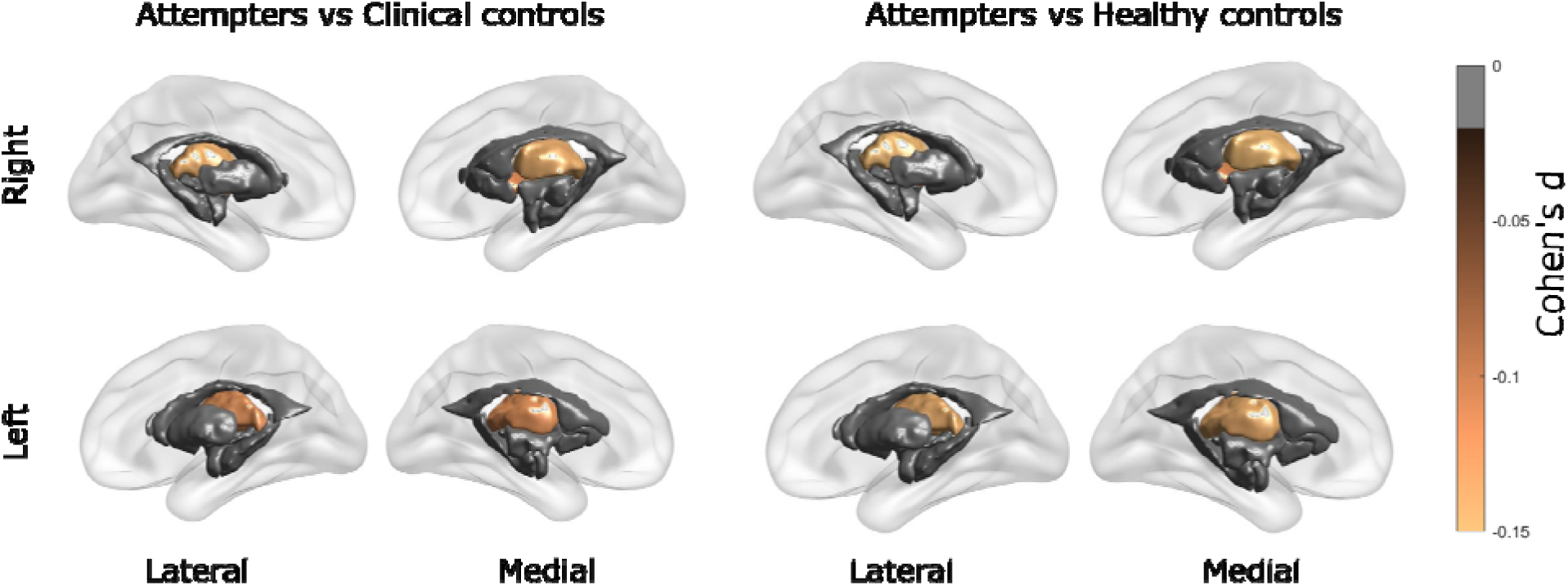
Group differences in subcortical volumes. Effect sizes are shown for regions that displayed a statistically significant difference in subcortical volumes between the groups: attempters compared to clinical controls (*left panel*) and attempters compared to healthy controls (*right panel*). No difference between clinical and healthy controls reached statistical significance after correction for multiple comparisons. Significant results are the bilateral thalamus and right pallidum.

**Table 2.** Main effects of group and *post hoc* analyses for subcortical volumes association results.

| Brain region | FDRp | SA | HC | CC | SAvsCC<br>Cohen's<br>d [p] | SAvsHC<br>Cohen's<br>d [p] | CCvsHC<br>Cohen's<br>d [p] |
| --- | --- | --- | --- | --- | --- | --- | --- |
| Left<br>thalamus | 0.027 | 682 | 12,343 | 5,500 | -0.13<br>[0.002**] | -0.14<br>[3.51E-04**] | 0.004<br>[0.792] |
| Right<br>thalamus | 0.007 | 685 | 12,338 | 5,494 | -0.14<br>[0.001**] | -0.16<br>[4.58E-05**] | 0.007<br>[0.684] |
| Right<br>pallidum | 0.036 | 686 | 12,325 | 5,471 | -0.12<br>[0.004**] | -0.11<br>[0.004**] | -0.015<br>[0.349] |
| ICV | 0.035 | 670 | 12,104 | 5,475 | 0.09<br>[0.028*] | -0.13<br>[0.001**] | -0.026<br>[0.112] |

### Cortical surface area

Eight of the 68 cortical regions under analysis displayed a significant group effect (FDR<0.05). These regions included the left and right pericalcarine, left and right cuneus, left inferior parietal, left rostral-middle frontal, right lingual, and right fusiform gyri. Depressed attempters exhibited, on average, a smaller surface area of the left cuneus, left inferior parietal, left rostral middle frontal and right pericalcarine regions, compared to healthy controls (**Table 3**). Furthermore, clinical controls exhibited smaller surface areas in the right pericalcarine and right fusiform gyri compared to healthy controls and clinical controls (**Table 3**). The left and right cuneus, left inferior parietal, right pericalcarine, and right lingual also exhibited nominally significant differences between attempters and clinical controls (p<0.05, uncorrected). After correcting for *post-hoc* multiple testing, only the left inferior parietal surface area was significantly different between attempters and clinical controls (Cohen’s d =-0.12; **Figure 2**).

**Table 3.** Main effects of group and *post hoc* analyses for cortical surface area association results.

| Gyrus | FDRp | SA | HC | CC | SAvsCC<br>Cohen's<br>d [p] | SAvsHC<br>Cohen's<br>d [p] | CCvsHC<br>Cohen's<br>d [p] |
| --- | --- | --- | --- | --- | --- | --- | --- |
| Right pericalcarine | 0.005 | 683 | 12335 | 5475 | -0.08<br>[0.044*] | -0.13<br>[0.001**] | -0.05<br>[0.001**] |
| Left inferior parietal | 0.017 | 679 | 12298 | 5484 | -0.12<br>[0.002**] | -0.15<br>[2<br>.30E-04**] | -0.02<br>[0.120] |
| Right fusiform | 0.017 | 674 | 12257 | 5473 | -0.02<br>[0.580] | -0.1<br>[0.013*] | -0.05<br>[0.001**] |
| Left rostral middle frontal | 0.03 | 685 | 12352 | 5507 | -0.06<br>[0.144] | -0.12<br>[0.003**] | -0.04<br>[0.013*] |
| Right lingual | 0.03 | 686 | 12388 | 5503 | -0.08<br>[0.043*] | -0.10<br>[0.010*] | -0.04<br>[0.018*] |
| Left cuneus | 0.04 | 682 | 12319 | 5492 | -0.10<br>[0.015*] | -0.13<br>[0.002**] | -0.02<br>[0.133] |
| Left pericalcarine | 0.04 | 683 | 12367 | 5500 | -0.07<br>[0.090] | -0.11<br>[0.008*] | -0.04<br>[0.019*] |
| Right cuneus | 0.05 | 682 | 12318 | 5465 | -0.08<br>[0.037*] | -0.10<br>[0.009*] | -0.03<br>[0.051] |

**Figure 2.**
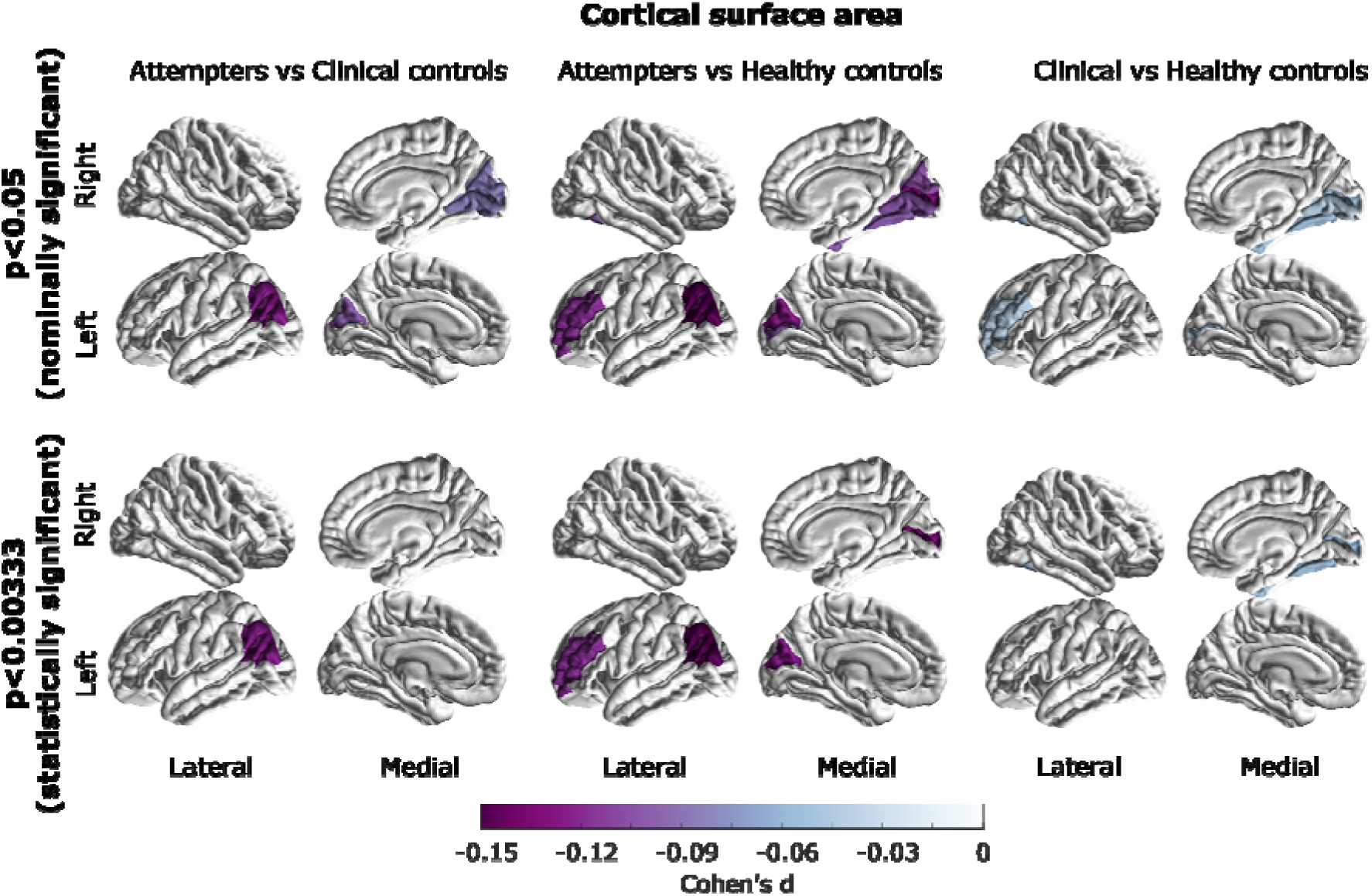
Group differences in cortical surface area. Effect sizes are shown for regions that displayed a (top) nominally significant (p<0.05) or (bottom) a statistically significant (p<0.00333 threshold after multiple test correction) posthoc difference between attempters and healthy controls (left panel), attempters and clinical controls (middle panel) and clinical controls compared to healthy controls (right panel). All of the colored regions showed a statistically significant group effect (FDR<0.05).

### Cortical thickness

Widespread cortical thickness differences between the three groups were observed (**Table 4**). Five of these regions showed a significant difference when comparing attempters to healthy controls, and three out of those five (left fusiform, left insula and left rostral middle frontal) showed nominally significant lower cortical thickness in depressed attempters compared to clinical controls. Only the left rostral middle frontal region displayed a statistically significant difference between attempters and healthy controls (**Table 4**). The left fusiform, and the left insula also showed a nominally significant difference between clinical and healthy controls. Conversely, the left rostral middle frontal did not show a significant difference between clinical and healthy controls. All regions with a nominally significant difference between attempters and clinical controls were in the left hemisphere. After adjusting for multiple testing, none of the cortical thickness differences between attempters and clinical controls reached statistical significance (**Figure 3**).

**Table 4.** Main effects of group and post-hoc results for cortical thickness

| Gyrus | FDRp | SA | HC | CC | SAvsCC<br>Cohen's<br>d [p] | SAvsH<br>C<br>Cohen's<br>d [p] | CCvsHC<br>Cohen's<br>d [p] |
| --- | --- | --- | --- | --- | --- | --- | --- |
| Left fusiform | 0.001 | 682 | 12350 | 5497 | -0.081<br>[0.047*] | -0.148<br>[1.78E-04**] | -0.054<br>[0.001**] |
| Left pars<br>opercularis | 0.001 | 684 | 12359 | 5498 | -0.079<br>[0.052] | -0.151<br>[1.22E-04**] | -0.057<br>[4.00E-04**] |
| Left rostral<br>anterior<br>cingulate | 0.001 | 687 | 12370 | 5494 | 0.011<br>[0.778] | -0.062<br>[0.112] | -0.066<br>[5.30E-05**] |
| Left insula | 0.002 | 687 | 12346 | 5499 | -0.098<br>[0.016*] | -0.135<br>[0.001**] | -0.047<br>[0.003*] |
| Right inferior<br>temporal | 0.004 | 677 | 12321 | 5478 | -0.070<br>[0.088] | -0.150<br>[1.44E-04**] | -0.043<br>[0.009*] |
| Right rostral<br>anterior<br>cingulate | 0.004 | 685 | 12356 | 5491 | -0.005<br>[0.906] | -0.054<br>[0.168] | -0.055<br>[0.001**] |
| Right fusiform | 0.004 | 685 | 12373 | 5502 | -0.009<br>[0.827] | -0.102<br>[0.009*] | -0.056<br>[0.001**] |
| Right insula | 0.004 | 687 | 12315 | 5492 | -0.049<br>[0.225] | -0.111<br>[0.005*] | -0.046<br>[0.005*] |
| Left posterior<br>cingulate | 0.007 | 683 | 12389 | 5511 | -0.008<br>[0.836] | -0.056<br>[0.153] | -0.050<br>[0.002*] |
| Left medial<br>orbitofrontal | 0.007 | 687 | 12338 | 5493 | -0.046<br>[0.255] | -0.096<br>[0.014*] | -0.038<br>[0.019*] |
| Left inferior temporal | 0.007 | 672 | 12314 | 5453 | -0.104<br>[0.011*] | -0.125<br>[0.002*] | -0.025<br>[0.131] |
| Right middle temporal | 0.008 | 684 | 12357 | 5483 | -0.053<br>[0.188] | -0.115<br>[0.003*] | -0.044<br>[0.006*] |
| Right superior temporal | 0.013 | 682 | 12283 | 5419 | -0.053<br>[0.196] | -0.124<br>[0.002*] | -0.044<br>[0.006*] |
| Right posterior cingulate | 0.018 | 685 | 12376 | 5518 | -0.019<br>[0.638] | -0.052<br>[0.187] | -0.042<br>[0.010*] |
| Right medial orbitofrontal | 0.02 | 686 | 12346 | 5504 | -0.046<br>[0.254] | -0.078<br>[0.047*] | -0.038<br>[0.020*] |
| Left rostral middle frontal | 0.02 | 682 | 12347 | 5497 | -0.088<br>[0.030*] | -0.132<br>[0.001**] | -0.019<br>[0.251] |
| Left superior frontal | 0.024 | 684 | 12343 | 5500 | -0.088<br>[0.031*] | -0.120<br>[0.002*] | -0.018<br>[0.258] |
| Left pars triangularis | 0.024 | 685 | 12349 | 5503 | -0.089<br>[0.028*] | -0.116<br>[0.003*] | -0.013<br>[0.41] |
| Right superior frontal | 0.037 | 685 | 12358 | 5503 | -0.047<br>[0.25] | -0.089<br>[0.024*] | -0.030<br>[0.067] |
| Left pars orbitalis | 0.042 | 686 | 12365 | 5505 | -0.098<br>[0.016*] | -0.116<br>[0.003*] | -0.002<br>[0.889] |
| Left caudal middle frontal | 0.042 | 683 | 12344 | 5504 | -0.122<br>[0.003*] | -0.116<br>[0.003*] | 0.013<br>[0.42] |
| Right isthmus cingulate | 0.048 | 686 | 12377 | 5506 | 0.007<br>[0.865] | -0.046<br>[0.241] | -0.036<br>[0.028*] |
| Right pars opercularis | 0.048 | 681 | 12357 | 5504 | -0.033<br>[0.415] | -0.107<br>[0.007*] | -0.030<br>[0.067] |

**Figure 3.**
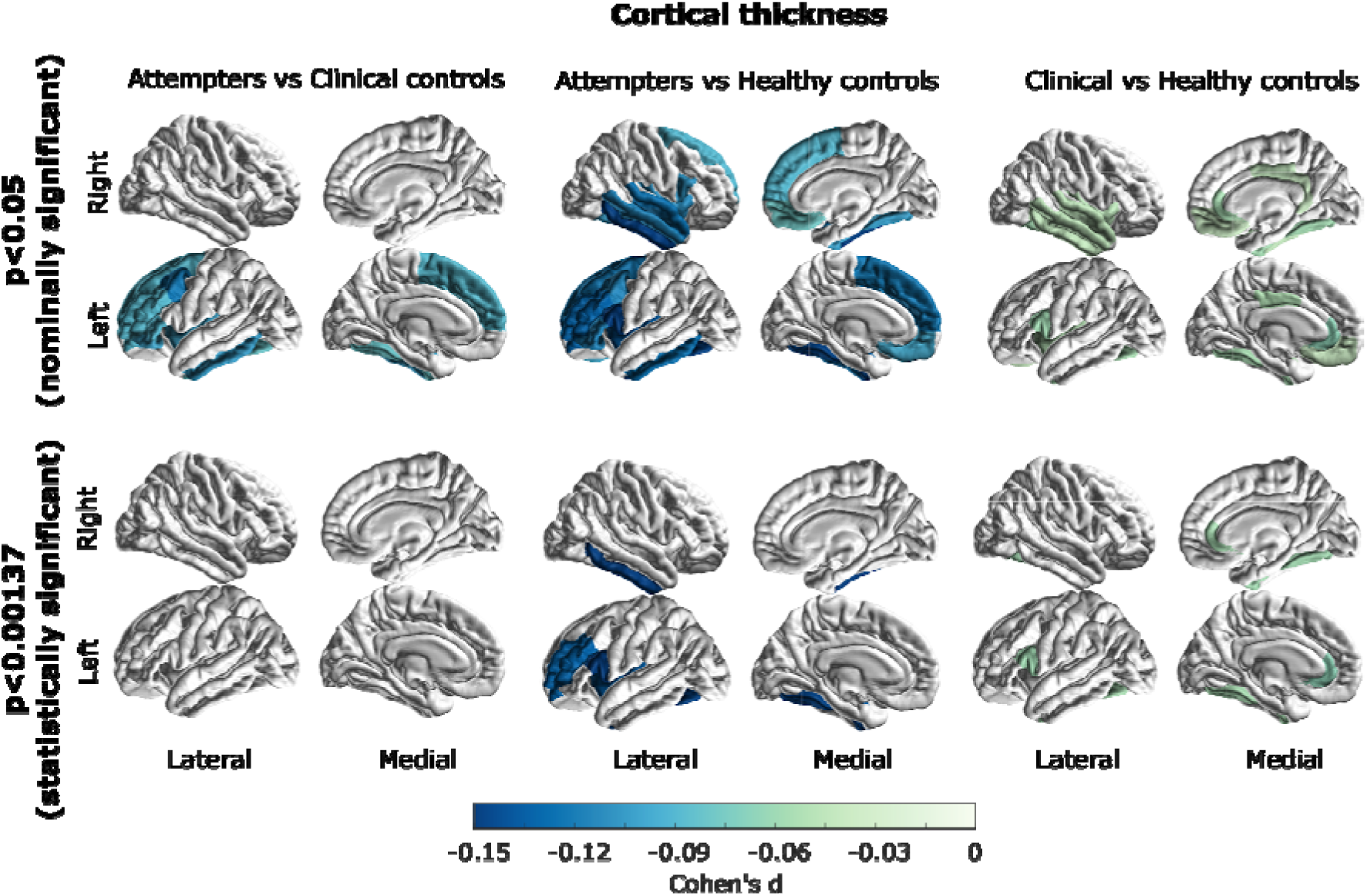
Group differences in cortical thickness. Effect sizes are shown for regions that displayed a (top) nominally significant (p<0.05) or (bottom) a statistically significant (p<0.001373 threshold after multiple test correction) posthoc difference between attempters and healthy controls (left panel), attempters and clinical controls (middle panel) and clinical controls compared to healthy controls (right panel). All of the colored regions showed a statistically significant group effect (FDR<0.05).

### Sensitivity analyses

Finally, we performed sensitivity analyses to assess the effect of additional covariates on the associations discovered. Namely, we tested whether our results were robust to adjustment for previous antidepressant use, depression severity, age of onset, and recurrence. These analyses had lower statistical power as data on these variables was available in fewer cohorts. Participants with a history of suicide attempt continued to show a smaller volume of the right thalamus (p<0.05) even after adjusting for history antidepressant use, depression severity, age of onset and recurrence (**Supplementary Tables S6-S9**). No other region remained significant after adjusting for any of these variables.

## DISCUSSION

The present study addressed the lack of statistical power, which may have resulted in low replicability and consistency in prior neuroimaging studies of suicide attempt. We pooled data across eighteen cohorts from around the world (n=18,925). We employed linear mixed models to test the association between history of suicide attempt and brain morphometry phenotypes while accounting for the site to site variation^34^. Our analyses revealed four regions associated explicitly with suicide attempt, above and beyond the effect of MDD, supporting the hypothesis of suicidality-associated neural differences^27^. Despite sample size, the relatively few regions identified, and the overall small effect sizes should be taken as a warning for future studies attempting to perform these types of analyses in small samples. We observed statistically significant volume reductions of the left and right thalamus, right pallidum, and a smaller surface area in the left inferior parietal cortex in depressed participants with a history of suicide attempt compared to both clinical and healthy controls. Prior studies have documented that these regions are associated with suicidal behaviors;^16,20,35–37^ however, the lack of evidence for associations in better-powered studies^11^ and the lack of consensus in the field^27^ made it challenging to reach definitive conclusions.

The pallidum has been linked to reward response, social activity mediation, and positive affect.^38,39^ Furthermore, a recent structural MRI study has linked the pallidum to suicidal ideation severity and impulsivity in a small sample of Korean MDD patients^40^. Thus alterations in the pallidum may reflect changes in affect and impulsivity known to be associated with suicidality.^41,42^ The thalamus, historically viewed as a passive gateway linking different brain regions, has been recently proposed to operate as an integrative hub of the brain^43^. A growing body of evidence suggests the different nuclei within the thalamus are involved in high order cognition^44^. Thalamic abnormalities and lesions have been linked to disorders such as addiction^45^, bipolar disorder,^46^ and schizophrenia.^47,48^ This is consistent with previously reported changes in the expression of glial fibrillary acidic protein (GFAP), a protein known to correlate with astrocyte function in the thalamus of subjects that died by suicide compared to matched controls^49^. Therefore, an impaired astrocytic function in the thalamus might mediate suicidality through impaired affect, empathy, and processing of stimuli-response relationships.

Our analyses of cortical regions identified several group differences in thickness and surface area that were associated with suicide attempt but did not survive strict multiple comparisons correction. Specifically, lower surface area in the right pericalcarine, left inferior parietal and left cuneus showed a statistically significant difference between attempters and healthy controls but only a nominally significant difference between attempters and clinical controls. The exception was the left inferior parietal lobe, which survived post-hoc multiple testing correction. As previously reviewed ^28^, the parietal region is part of the executive control network, which exerts control over thought, emotion and behavior, and has been linked to suicide attempt in bipolar disorder ^50^

Conversely, we observed no robust cortical thickness differences associated with a history of suicide attempt. Some regions differed between attempters and healthy controls, but the lack of differences between clinical controls and suicide attempters suggests that these associations may be driven by depression status, rather than suicidality. The fact that subcortical volume and surface area associations had a stronger association with suicide attempt than with cortical thickness could be of interest in light of genetic analyses that identified substantial differences in the genetic etiology of cortical morphometry phenotypes ^51–53^ For example, surface area measurements were reported to have a higher heritability and be more influenced by early developmental genetic influences compared to cortical thickness,^52^ suggesting that biomarkers associated with surface area are likely established earlier in life. In contrast, cortical thickness phenotypes may be more variable and more susceptible to adverse environmental effects later in life, such as substance use or having a psychiatric condition. Future studies could test this hypothesis through longitudinal analyses of suicidality risk and brain morphometry.

The substantial overlap between depression severity and suicidality (e.g., our Table 1 shows that participants with a past suicide attempt also have higher HDRS and Beck Depression Inventory (BDI) sum scores even after removing the suicide item) makes it hard to differentiate whether an identified statistical effect is driven by depression severity or suicide attempt status. This is also supported by the fact that several cortical associations also displayed an effect when comparing clinical to healthy controls, and that those results did not remain significant after correcting for multiple comparisons in most of our sensitivity analyses that controlled for proxies of depression severity. Overall, effect sizes were smaller when comparing clinical to healthy controls than those when comparing suicide attempters to either control group. That might be due to the previously discussed collinearity between suicidality and depression severity. Many of the most severely depressed cases will be among the depressed suicidal group, thus reducing the strength of any signals associated with depression severity.

Our previous work only assessed subcortical correlates of suicidality and did not show any significant associations in a smaller sample^11^. Notably, that study identified nominally significant associations of suicidality with a smaller intracranial and thalamic volumes, but the association did not survive multiple testing corrections. Our current intracranial and thalamus volumes results are consistent with our previous observations. In addition to the significantly increased statistical power in the current study, there are significant design differences between our previous study and the current investigation. First, the implementation of a mega-analytical framework instead of a meta-analytical approach enabled us to include more samples with a relatively small number of suicide attempt cases. Accordingly, we implemented a linear mixed-model framework, which allowed us to adjust for the potential differences across sites by modeling imaging-site as a random effect without significantly compromising statistical power. This represents a substantially more comprehensive study, as it includes cortical measures in addition to subcortical and ventricular volumes. Finally, the previous study focused on any suicidal symptoms (including suicide ideation, suicide planning, and history of suicide attempt) while here we focused solely on suicide attempt; thus, avoiding the heterogeneity arising from defining and including other forms of suicidality.

This study examined cohorts from the ENIGMA-MDD working group and a subset of participants fulfilling MDD diagnosis criteria from the UK Biobank. Therefore, all individuals with a history of suicide attempt also had an MDD diagnosis. A limitation of this approach is that we cannot conclude whether the suicidality associations observed in this study would also be correlated with suicide attempt history in other mental illnesses. Only the right thalamus was associated with suicide attempt above and beyond potential covariates such as depression severity, recurrence, and history of antidepressant use. The observation of the left thalamus, right pallidum, and left inferior parietal cortex surface area no longer showing an association with suicide attempt after correcting for these additional clinical variables was expected for several reasons. First, suicidality is highly collinear with depression severity, including recurrence of depressive episodes (see **Table 1**). Second, the lack of information on these variables in some of the contributing cohorts greatly reduced the sample size for the sensitivity analyses, resulting in reduced statistical power to detect the same small effect sizes that we observed for these brain measures across the whole sample. Future collaborative efforts would benefit from a cross-disorder design comparing suicidality and its correlates across different conditions.

Overall, this international collaboration has yielded valuable insights into the neurobiology of suicide. The number of associations discovered, and their small effect sizes indicates that morphological differences between attempters and non-attempters will likely not be useful for clinical diagnosis or risk stratification. Even so, if several neural circuits underlie suicidality with relatively small effect sizes, the aggregation of them might still be useful for risk stratification. That is beginning to become apparent in the field of genetics, where the liability to complex traits can only be modeled through the integration of hundreds to thousands of genetic variants, each with a small effect size. If this is the case, increasing the resolution of neuroimaging studies (e.g., to the vertex level) or integrating structural measurements with other modalities (e.g., functional imaging data), will help.

In summary, our results suggest that suicide attempt is associated with volumetric reductions within the thalamus, right pallidum and surface area reductions in the left interior parietal lobe, over and above the effects of depression alone. Our findings suggest that several regions are associated with suicide attempt, albeit with relatively small effect sizes. This study addressed the lack of replicability and consistency in several previously published neuroimaging studies of suicide attempt and further demonstrated the need for well-powered samples and collaborative efforts to avoid reaching biased or misleading conclusions. Future neuroimaging studies of suicidality should focus on: achieving even larger sample sizes, studying brain images at a finer resolution and using other MRI modalities, and investigating the neurological underpinnings of suicidal behavior across individuals with different diagnoses.

## Data Availability

Data and code requests will be evaluated on a case by case basis to comply with IRB policies and regulations. Interested researchers may contact the corresponding author of this study for information.

## ACKNOWLEDGMENTS

The work reported here was supported in part by many public and private agencies across the world. **Core** funding for ENIGMA was provided by the NIH Big Data to Knowledge (BD2K) program under consortium grant U54 EB020403. **AFFDIS** was supported by the University Medical Center Göttingen and the German Federal Ministry of Education and Research (Bundesministerium fuer Bildung und Forschung, BMBF: 01 ZX 1507, ‘‘PreNeSt - e:Med’’). The **BiDirect** study was supported by grants from the German Federal Ministry of Education and Research (BMBF; grants FKZ-01ER0816 and FKZ-01ER1506). **ETPB** study at NIH is funded by the National Institute of Mental Health Intramural Program. The **FOR2107** study was supported by the German Research Foundation (DFG) grant Nos Ki588/14-1 and Ki588/14-2 to TK. The **IMH Singapore** cohort was supported by the National Healthcare Group Research Grant (SIG/15012) awarded to KS. The **LOND** study was supported in part by the Biomedical Research Centre, South London and Maudsley NHS Foundation Trust, London, UK. The **Stanford** study was supported in part by the National Institute of Mental Health Grant R37-MH101495 to IHG. The **Melbourne** study was supported by Australia’s National Health and Medical Research Council (NHMRC) Project Grants 1064643 (principal investigator, BJH) and 1024570 (principal investigator, CGD). The **QTIM** study was supported by the National Institute of Child Health and Human Development (R01-HD050735), and the National Health and Medical Research Council (NHMRC 486682, 1009064), Australia (principal investigator MJW). The **Pharmo** study in Amsterdam was supported by faculty resources of the Academic Medical Center, University of Amsterdam, and 11.32050.26 ERA-NET PRIOMEDCHILD FP 6 (EU). The **UCSF** study was funded in part by the National Institute of Mental Health (R01MH085734 and K01MH117442), National Center for Complementary and Integrative Health (NCCIH 1R61AT009864) and the American Foundation for Suicide Prevention (AFSP). The **Edinburgh** group is funded by a Wellcome Trust Strategic Award “Stratifying Resilience and Depression Longitudinally” (STRADL) (Reference 104036/Z/14/Z). The **SHIP** cohort is part of the Community Medicine Research Network of the University Medicine Greifswald, which is supported by the German Federal State of Mecklenburg-West Pomerania. AIC is supported by a UQ Research Training Scholarship from The University of Queensland (UQ). MER thanks the support of the NHMRC and Australian Research Council (ARC) through a Research Fellowship (APP1102821). BJH and CGD were supported by NHMRC Career Development Fellowships (1124472 and 1061757, respectively). SEM thanks funding support from NHMRC grants APP1103623, APP1158127, and APP1172917. TTY is funded in part by NCCIH 1R61AT009864. LS and NJ received support from the National Institute of Mental Health of the National Institutes of Health (USA) under Award Number R01-MH117601.

## AUTHOR CONTRIBUTIONS

MER and LS conceived and jointly supervised the study. Authors from all site cohorts were involved in data collection, processing, analysis, and funding for their samples. AIC implemented the analysis pipeline, performed all statistical analyses, and generated the tables and figures. AIC, MER, and LS drafted the first draft of the manuscript with feedback and input from all co-authors. All authors approved the content of this manuscript.

## COMPETING INTERESTS

Hans J. Grabe has received travel grants and speakers’ honoraria from Fresenius Medical Care, Neuraxpharm, Servier, and Janssen Cilag as well as research funding from Fresenius Medical Care; Carlos A. Zarate is a full-time U.S. government employee. He is listed as a co-inventor on a patent for the use of ketamine and its metabolites in major depression and suicidal ideation. Dr. Zarate has assigned his patent rights to the U.S. government but will share a percentage of any royalties that may be received by the government. NJ and PMT are MPI of a research related grant from Biogen, Inc., for work unrelated to this manuscript.

